# Impact assessment of social behavioral change activities on infant and young child feeding (IYCF) of the nutrition program in the host community

**DOI:** 10.1101/2025.11.17.25340405

**Authors:** Taslima Arzu, S.M. Symon Bappy, Md. Alamgir Hossain, Monowarul Islam, Vulon Prosad, Suparna Das Toma, Md. Ariful Kabir Sujan

## Abstract

**Aims:** The purposes of the impact assessment for the Social Behavioral Change (SBC) activities in the nutrition program in the host community were: to measure the impact of the Social Behavioral Change (SBC) activities to ensure the optimum infant and young child feeding (IYCF), especially on the pregnant and caregiver of under 2 years children on IYCF activities and to provide recommendations based on an overall assessment.

**Study design:** The study was designed to review the secondary nutrition program performance data and conduct a cross-sectional study using both quantitative and qualitative data collection to gain deep insight into the impact of the SBC activities on changing the behavior of the target population.

**Place and Duration of Study:** Data was collected in the host community, where the coastal & the poorest people from the host community were staying and getting nutrition support in Moheshkhali and Pekua of Coxs Bazar district of Bangladesh.

**Methodology:** A household survey was conducted to collect quantitative data from the project beneficiaries. This survey was tailored to gather relevant indicators and assess the effectiveness of the interventions. Quantitative data were collected from the beneficiaries through household surveys. We included 224 pregnant and lactating women (PLW) and 227 caregivers of 0-59-month-old children (4 males and 447 females). Qualitative data were obtained through Focus Group Discussions (FGDs-8; 8-10 females) and Key Informant Interviews (KIIs-8; MtMSG members, nutrition and health staff, and participants from cooking demonstrations).

**Results:** A complete package of SBC approach to breaking the social stigma and barrier targeting the audience through different methods; for example, to improve the Infants and Young Child Feeding (IYCF) indicators by providing IYCF messages through health and nutrition education, group messaging at IYCF corner, Community sensitization, meetings and workshops, mother-to-mother support groups, and IYCF counseling from health facilities. Complementary feeding cooking demonstration for hands-on learning can change the negative behavior of the targeted audience in a positive direction. In this study, we found that 61.2% of mothers changed one of the negative behaviors related to IYCF in a positive direction.

All mothers knew the importance of exclusive breastfeeding, and 73.0% of mothers practiced it properly, which is higher than the current exclusive breastfeeding rate of 62.1% (IYCF Survey 2022 by ACF) (11). 52.8% of mothers ensured the minimum dietary diversity, and 35.8% ensured the Minimum Acceptable Diet (MAD) of complementary feeding for children 6-23 months, which came from the complete SBC approach to IYCF practices.

**Conclusion:** This comprehensive study of secondary data review and qualitative findings with validation through the quantitative survey finds that a comprehensive nutrition activity engaging the target population in the SBC activities changes the social stigma and barrier.

All mothers who attended the study expressed proper knowledge of IYCF and also emphasized their physical and mental health to ensure proper IYCF practices for children 0-23 months, such as exclusive breastfeeding for the first six months, starting complementary feeding after six months, and ensuring the minimum food diversity to ensure proper nutrition of the young children. This group of mothers can work and support other mothers in the best practices of IYCF and break the social stigma and barriers of IYCF practices.

## 1. Introduction

As the community-based management of acute Malnutrition (CMAM) nutrition program has been running since 2014 in the host community of Moheshkhali and Pekua of Cox’s Bazar district, the malnutrition situation remains critical (Global Acute Malnutrition, GAM is 14.7% in Moheshkhali and 11.7% in Pekua at the year of 2021(1, 2), especially for children 6-23 months of 20.0%. Early childhood malnutrition leads to chronic malnutrition, such as stunting among older children (Stunting among children 6-23 months; 27.5% & 24-59 months; 34.4% (2) in Moheshkhali and among children 6-23 months; 27.5% & 24-59 months; 34.4% in Pekua (1). We are focusing here on the SBC approach to improve malnutrition and negative behaviour, which are affecting children under 2 years with a high Global Acute Malnutrition (GAM) rate in a positive direction, so that the malnutrition situation can improve and the sustainability of the nutrition messages.

Malnutrition is a persistent health problem among children in Bangladesh, especially children under 2 years, due to the lack of proper weaning foods, both diverse and balanced (3,4). The optimal infant and young child feeding practices during the first 2 years of life are of paramount importance as this period is the “critical window” for the promotion of health, good growth, and behavioral and cognitive development. Optimal infant and young child feeding practices include initiation of breastfeeding within 1 hour of birth, exclusive breastfeeding for the first 6 months, and continuation of breastfeeding for 2 years or more, along with nutritionally adequate, safe, age-appropriate, responsive complementary feeding starting at 6 months. [5]. It was estimated that about one-fifth of overall under-five mortality can be averted if 90% of infants are covered with an inclusive package of interventions to promote, protect, and support the optimal infant and young child feeding (IYCF) practices. [6]. A large proportion of children become vulnerable to stunting, poor cognitive development, and significantly increased risk of infectious diseases, such as diarrhea and acute respiratory infection, due to poor complementary feeding practices. [7]. Epidemiological evidence of a causal association between early initiation of breastfeeding and reduced infection-specific neonatal mortality has also been documented. [8]

The findings from the 2022 IYCF survey among the host communities offer valuable insights, highlighting both encouraging trends and areas of concern. While 62.1% of children under six months were reported to be exclusively breastfed, the continued practice of giving prelacteal feeds—often driven by cultural and religious beliefs—remains an issue (9). Among children aged 6–23 months, only 31.3% met the Minimum Dietary Diversity (MDD) and 26.0% met the Minimum Acceptable Diet (MAD) criteria, which is particularly alarming (9). The positive aspect is that the caregiver engagement in nutrition education is notably high, with 96.0% reporting active participation in sessions over the past 23 months, despite generally low education levels among caregivers of children aged 0–23 months. Only 4.0% reported being unable to attend these sessions during the same period. (10)

The optimum IYCF practices have a great impact on the physical and mental development of the child (11). Breastfeeding strengthens emotional security and affection, creating a strong bond between the mother and the child, which in turn promotes the psychosocial development of a child. To ensure the good nutrition status of the infant as well as the mother, maternal nutrition plays a vital role. Breastfeeding is nature’s way of nurturing the child. It provides learning and development opportunities for the infant. Breast milk also leads to increased intelligence quotients and better visual acuity due to the presence of special fatty acids in it. (12)

The IYCF practices are strongly influenced by what people recognize, think, and believe, and who traditionally encourage mothers to breastfeed by giving information on the benefits of breastfeeding for the infant and the mother herself. Women’s behaviour can be easily positively modified through religious teachings. Breastfeeding may be affected by religious ideologies using the doctrine in religious texts, affected by social circumstances, and economic factors. Effective communication for behavioural change is necessary for ensuring optimal infant feeding (11,13,14). Counselling the mothers by reinforcing the cultural and religious practices supporting breastfeeding can help enormously. The use of local religious teachings can bring positive changes in the implementation of health & nutrition programs (11,12). Community-based IYCF counselling and support can play an important role in improving these practices: it can ensure access to these services in the poorest and the most vulnerable communities with limited access to health care and therefore become an important strategy for programming with an equity focus. (11,12). Public nutrition education that promotes infant and young child feeding as defined by WHO, considering social-cultural factors, is needed and recommended (11,15).

The social behavioural change (SBC) communication activities include community screening of children and pregnant and lactating women (PLW) with the dissemination of IYCF messaging at the household level, courtyard sessions with complementary cooking demonstration, Growth monitoring, and promotion (GMP) of the children under five years. The optimum maternal, infant, and young child feeding (IYCF) and care practices through one-to-one IYCF need-based messaging, group messaging, Mother to Mother Support Groups (MtMSG), Father Support Group, and community sensitization through World Breastfeeding Week celebration, Nutrition Action Week, etc.

The purposes of the impact assessment for the SBC activities in the nutrition program in the host community were: to measure the impact of the Social Behavioural Change (SBC) activities to ensure the optimum infant and young child feeding (IYCF), especially on the pregnant and caregiver of under 2 years’ children on IYCF activities and to provide recommendations based on an overall assessment to improve the IYCF conditions for better outcome of the nutrition program.

## 2. Method And Materials

The tools used both qualitative and quantitative methods to ensure a thorough assessment by cross-checking data.

### 2.1. Sampling Frame

Households and individual beneficiaries from the Moheshkhali and Pekua were the focus of our data collection efforts.

### 2.2. Quantitative Data

A household survey was conducted to collect quantitative data from the project beneficiaries. This survey was tailored to gather relevant indicators and assess the effectiveness of the interventions. Quantitative data were collected from the beneficiaries by a household survey. The sample size calculator Raosoft (where confidence level is 95%, response distribution is 50%, and margin of error is 6.5%) was used to calculate the sample size for the Host Community.

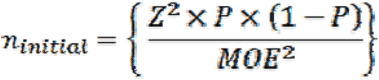

Where,

- Initial = minimum required initial sample size (before adjusting for finite population correction).
- p = an estimate of the true (but unknown) population (project participant) proportion at baseline=50% (0.50).
- Z = critical value from a normal probability distribution (Z-score corresponding to the 95% confidence level) [Z = 1.96 at 95% confidence level].
- MOE = margin of error (acceptable percentage error) = 0.065 (6.5%).
- d = Design effect, a two-stage PPS cluster sampling procedure is proposed, and in that case, the design effect might be close to two [d = 2].
- NF = non-response factor [1.10] (assumes a 10% non-response rate).

**Table 1:**
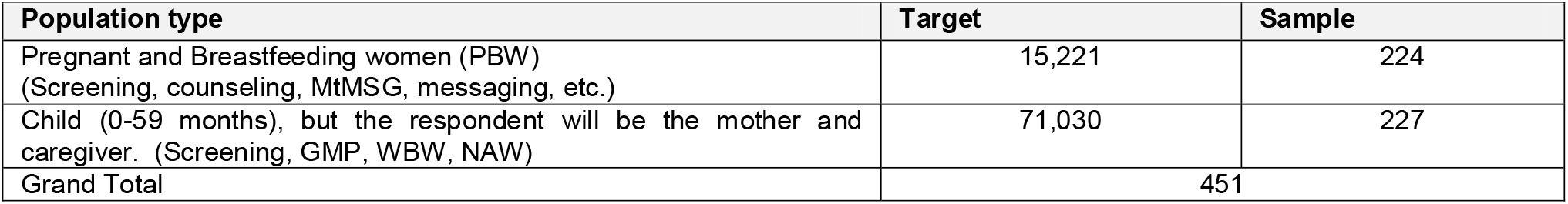
Population type and quantitative sample for the assessment.

### 2.3. Qualitative Data: Qualitative data were obtained through Focus Group Discussions (FGDs) and Key Informant Interviews (KIIs)

**Table 2:**
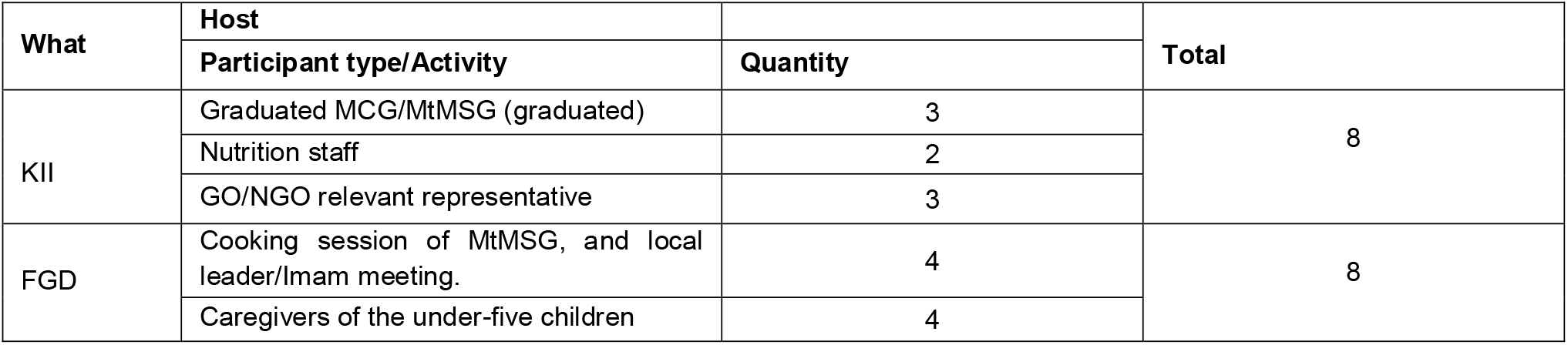
Number of FGD and KII conducted.

| What | Host | Quantity | Total |
| --- | --- | --- | --- |
|  | Participant type/Activity |  |  |
| KII | Graduated MCG/MtMSG (graduated) | 3 | 8 |
|  | Nutrition staff | 2 |  |
|  | GO/NGO relevant representative | 3 |  |
| FGD | Cooking session of MtMSG, and local leader/Imam meeting. | 4 | 8 |
|  | Caregivers of the under-five children | 4 |  |

### 2.4. Data Collection and Capacity Building

Before the start of data collection for the host community, the nutrition staff and the Monitoring, Evaluation, Accountability and Learning (MEAL) team served as data collectors. The MEAL team conducted orientation sessions for both groups on the proper use of all tools, with support from the Deputy Program Manager and Program Manager if needed.

During the orientation, the program team provided the venue and snacks for the data collection team. The team conducted a pilot test of the finalized tools to ensure they functioned as expected and captured the required data. All team members are proficient in both English and the local language, and have experience using the KoBo Toolbox or similar ODK platforms. Data collection began once the team had demonstrated full confidence and competence with the tools and processes.

### 2.5. Ensuring Reliability and Validity

To maintain high data quality throughout the process, it was ensured that all the data collection tools were in English and understandable to the team, and all enumerators were provided with thorough training before data collection began, ensuring they were proficient in communicating in the local language. The data collection tools were piloted and refined based on the feedback from the piloting phase. Continuous supervision was maintained throughout the data collection process to ensure accuracy and adherence to protocol. Regular reviews of the collected data were conducted, and corrective actions were taken as necessary. Data validation checks were performed to minimize errors; the datasheet and the data were error-free. Unnecessary or irrelevant questions were avoided in the tools, as this would increase data collection time for both qualitative and quantitative assessments.

### 2.6. Limitations

During the rainy season, heavy rainfall makes it uncomfortable for beneficiaries to participate in interviews, as they could get soaked or feel uneasy. To solve this, interviewers arranged safe and dry places where people can comfortably answer questions. Some women also felt shy or hesitant to share information due to cultural reasons. To overcome this, we chose interviewers (provided female Community Nutrition Volunteers (CNV) or Staff) who were sensitive to these issues and used respectful, culturally appropriate language in the questions. Additionally, political tensions or sudden weather changes, like storms and internet disruptions, could interrupt data collection. The team kept track of security updates and adjusted plans to ensure safety.

## 3. Result

### 3.1. Nutrition program coverage for the SBC activities

**Table 3:**
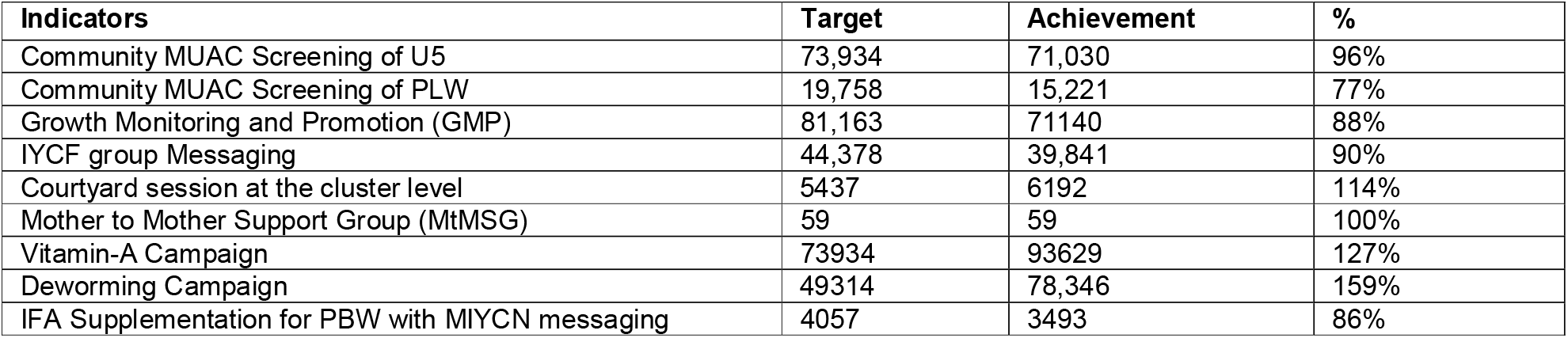
Program performance assessment by evaluating the target vs achievement.

### 3.2. Survey Results & Discussion

**Figure 1:**
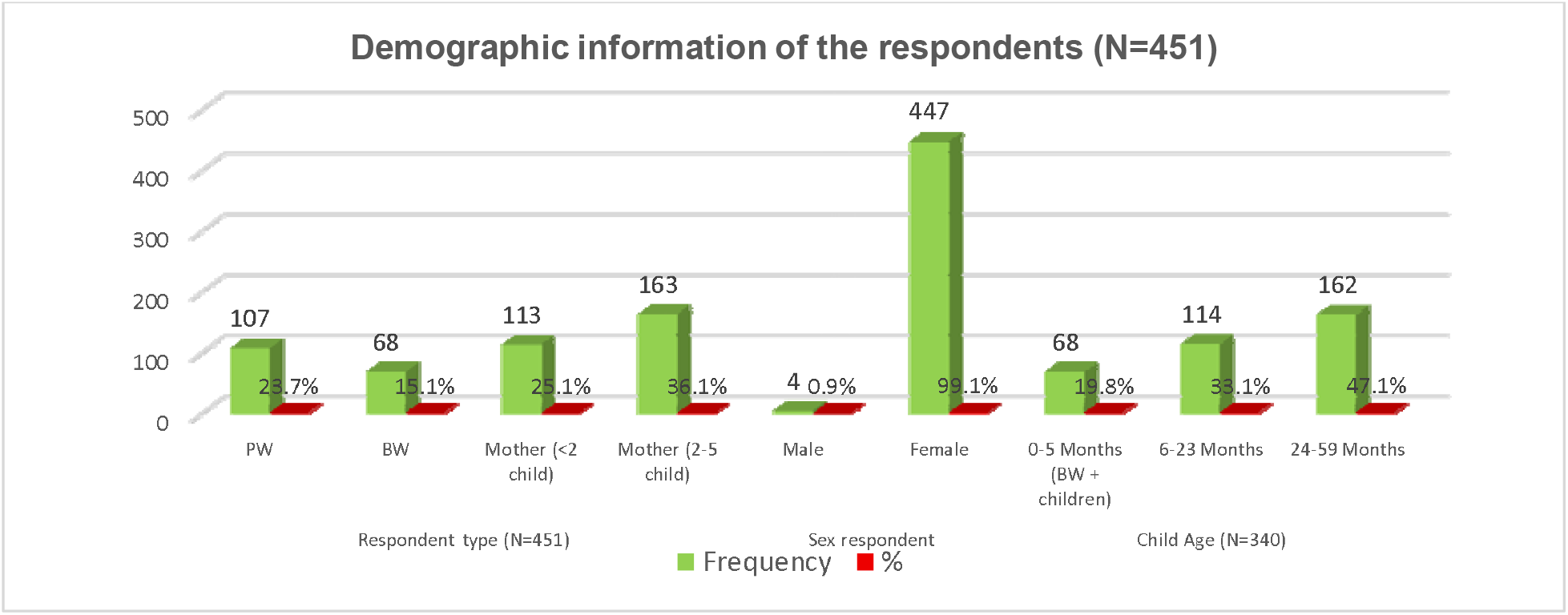
Demographic information of the respondents (N=451)

**Table 4:**
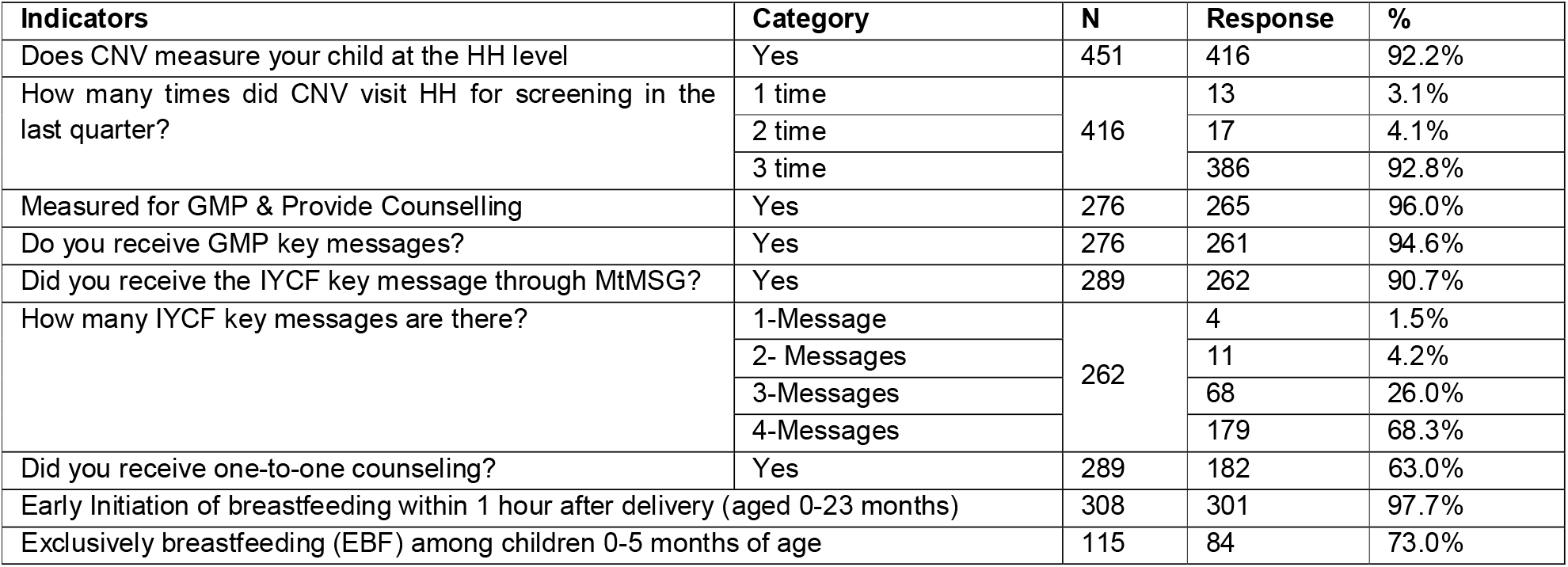
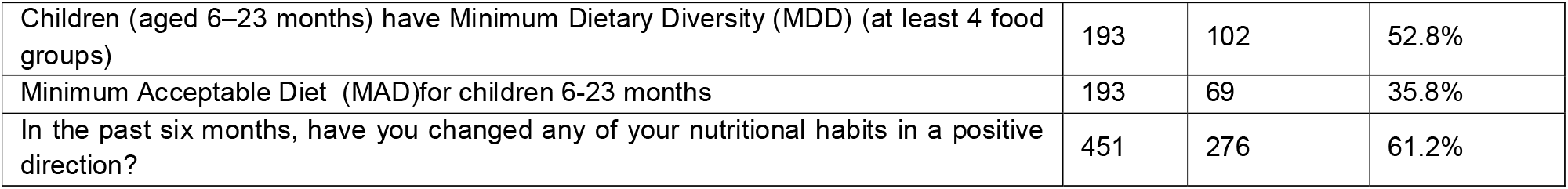
Results from the quantitative assessment.

### 3.3. Nutritional status of the children from the Survey

**Figure 2:**
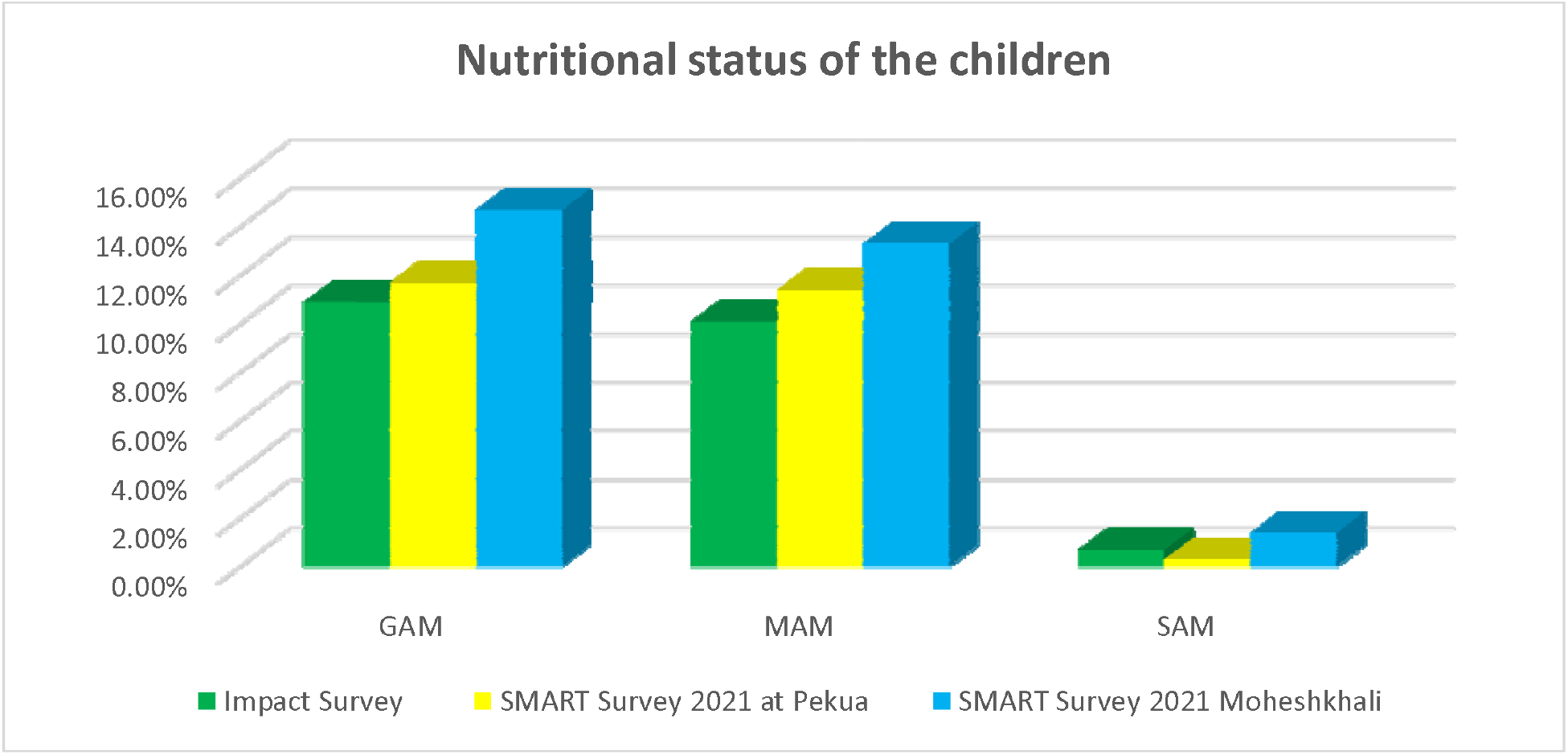
Nutritional status of the children whose caregivers or mothers attended SBC activities (Wasting GAM: Global Acute Malnutrition, SAM: Severe Acute Malnutrition, MAM: Moderate Acute Malnutrition, SENS: Standardized Extended Nutrition Survey

### 3.4. The result from the qualitative assessment

#### 3.4.1.1. Insights of KII (health and nutrition Staff)

- Areas with high malnutrition and vulnerability were chosen to help those who need it most by regular analysis of program data, program coverage & performance.
- Field visits and reports were used to check the work and ensure it was done well.
- Challenges like hard-to-reach areas and limited access for marginalized groups make it harder to continue the work long-term.
- Children under 2 years and pregnant or breastfeeding women were prioritized to get help first and ensure their visit to the IYCF corner for initial assessment of breastfeeding problems, and provide services according to the assessment findings.
- Communities were involved through activities like workshops and sensitization meetings to make them feel part of the project.
- Pregnant and lactating women are trained on IYCF, childcare, and feeding for the sustainability of the program activities through group messaging, counseling, and Mother to Mother Support group activities.

#### 3.4.1.2. Insights of KII and FGD

- Model mothers share key messages on breastfeeding, nutrition, and hygiene, improving child health, and empowering mothers in the community.
- A supplementary cooking session on complementary feeding (CF) with six female participants (one caregiver and five other women) improved understanding of breastfeeding, complementary foods, and hygiene practices, promoting better family health.
- A complementary cooking session with ten female participants (one PBW, nine caregivers) emphasized nutrient-rich feeding alongside breastfeeding, enhancing child nutrition and hygiene.
- A courtyard session with 14 participants (one male, 13 females: one PBW, five caregivers, eight other women) improved knowledge of child nutrition and hygiene, fostering healthier practices and active community sharing.
- 100% of lead mothers understand that the mother aims to enhance physical & mental health and well-being, and they also emphasize breastfeeding.
- 100% of participants from MtMSG mothers (Out of 11) knew that only the first breast milk for the first 6 months of the child.
- 100% of mothers’ positive response after 6 months of starting to feed complementary food.

## 4. Discussion

This comprehensive study of secondary data review and qualitative findings, with validation through the quantitative survey, finds that a comprehensive nutrition activity and engaging the target population in nutrition program activities change the social stigma and barriers & improve the nutritional status of the children and Pregnant and Breastfeeding women. A complete package of Social Behavioral Change (SBC) activities targeting the most vulnerable groups to break the malnutrition life-cycle through different methods; for example, to improve the nutritional status through improving the Infants and Young Child Feeding (IYCF) indicators by providing IYCF messages through health and nutrition education, group messaging at IYCF Community Clinic, Community sensitization, meetings and workshops, mother-to-mother support groups, IYCF counseling from health facilities, improving immunity through micronutrient (IFA, Vitamin-A, and Deworming, etc.) supplementation with IYCF messaging. The quantitative survey showed that the nutritional status (wasting) of the children improved from 11.7% in Pekua and 14.7% in Moheshkhali (1, 2) to 10.9% in the host community of Moheshkhali and Pekua.

The IYCF practices are strongly influenced by what people know, think, and believe, and are also affected by social circumstances and economic factors. Effective communication for behavioral change is necessary for ensuring optimal infant and young child feeding. Awareness regarding IYCF practices and their benefits in Maternal and Child Health (MCH). (4,11,12,13)

All mothers practiced the Infants and Young Child Feeding (IYCF) indicators; the importance of exclusive breastfeeding, and 73.1% of the mothers from the host community practice exclusive breastfeeding (62.1% 2022) (9). 35.8% of mothers from the host community ensured the Minimum Acceptable Diet (MAD), with 52.8% having the Minimum Acceptable Diet (MDD) of complementary feeding for children 6-23 months, which came from the complete SBC approach to IYCF practices.

Cooking demonstrations with proper messages on complementary feeding for children 6-23 months for hands-on learning can change the negative behavior of the targeted audience in a positive direction. In this study, we found that 61.1% of mothers from the host community changed one of the negative behaviors related to IYCF in a positive direction.

The IYCF practices are strongly influenced by what people know, think, and believe, and are also affected by social circumstances and economic factors. Effective communication for behavioral change is necessary for ensuring optimal infant and young child feeding. Awareness regarding IYCF practices and their benefits in Maternal and Child Health (MCH). (16,17,18, 19,20)

## 5. Conclusion & Recommendations

Despite these successes, challenges are not at an end: deep-rooted cultural and geographical isolation and economic constraints create unequal access to services. Among the recommendations that have emerged toward improving the impact and sustainability of the SBC approaches for outreach to marginalized or underrepresented groups with effective, culturally sensitive strategies and promotion of awareness through digital instruments. It is also critically important that male family members and local leaders be involved in community-level SBC campaigns.

Recommendations came from the impact assessment of SBC approaches:

- Address barriers faced by marginalized groups through targeted outreach, culturally sensitive materials, and ensuring accessible session locations and timings.
- Engage local leaders, model mothers, and community members in planning and implementing programs to foster ownership and trust, ensuring sustained impact.
- Increase the frequency of monitoring visits, conduct regular data analysis, and adapt interventions to align with evolving community needs and feedback.
- Utilize visual aids, hands-on demonstrations, and affordable, locally available food options to address affordability and improve message retention.
- Conduct regular training sessions for nutrition staff and volunteers, focusing on technical skills and service delivery.
- Scale up awareness campaigns focusing on practical, low-cost IYCF practices, emphasizing exclusive breastfeeding, complementary feeding, and hygiene as critical components of child health.
- Develop culturally appropriate messages to address barriers to breastfeeding and dietary diversity.
- Engage male family members and community influencers to support women’s participation in nutrition programs.

## Supporting information

Ethical Permission from the ethical authority

Population type and quantitative sample for the assessment

Number of FGD and KII conducted

Program performance assessment by evaluating the target vs achievement

Demographic information of the respondents (N=451)

Results from the quantitative assessment

Nutritional status of the children

## Data Availability

All data reserved to the corresponding author and if require, corresponding author will send the data to the publisher

## 6. Acknowledgement

On behalf of the Research team, we would like to acknowledge all Caregivers of children under five, Pregnant and Lactating Women, and the religious leaders who have shared their opinions and experiences in this study. Their ideas and suggestions are invaluable in future programming aiming to reduce the impact of ration cuts on the nutritional status of children under five, pregnant and lactating women, and other vulnerable groups.

We acknowledge the support of other stakeholders, including Nutrition staff who conducted qualitative studies through focus group discussions (FGDs) and key informant interviews (KIIs) at the field level.

## 7. Ethical Consideration

To protect the participants, especially children and pregnant or lactating women, the guiding principles for data collection included voluntarism, confidentiality, and anonymity. Participation was strictly voluntary. In the beginning, the purpose of the study was explained, and it was made certain that the information provided would be kept confidential. They were free to withdraw or skip any question without any repercussions.

### Do No Harm

The questions were constructed to do no harm and cause no discomfort to the respondents. Support was given for sensitive issues, or further resources were provided where appropriate.

### Integrity

Data reported fully and with accuracy. Full effort was made to double-check information and make sure information was presented correctly in its proper context.

### Participant Feedback

Findings related to key issues were fed back to the community and participants to incorporate their views into the final report.

### Child Protection

In sessions involving children, a responsible adult was also present; children were never asked anything that could make them remember something personal and hurt them. Interviewers followed guidelines for child protection strictly because it is relevant to safety.

## 8. Informed Consent

Before the start of quantitative and qualitative data collection, the survey team will have a minute or two of introduction and the purpose of the survey, and how long the survey will take for the respondents. The team will also guarantee the respondent or the FGD group the confidentiality and privacy of the information that will be collected during the survey. No personal and family information shall be revealed during reporting, and the rights and privacy of the respondents shall be respected. If she/he wishes not to respond to questions or wishes to drop the survey, both quantitative and qualitative. Therefore, the survey only proceeds upon getting informed consent from the respondent and or the FGD group participants.

The participants were selected equitably, and their informed consent was sought to ensure that they participated in the study voluntarily.

## 9. Ethical Approval

As the ethical approval authority at the district level for health and nutrition is the Civil Surgeon, approval for the survey was obtained from the respective authorities, including the Civil Surgeon (CS), Upazila Health and Family Planning Officer (UH&FPO), and the Chairman of the respective Unions.

## 10. Supporting Information

S1 Table 1: Population type and quantitative sample for the assessment (PDF)

S2 Table 2: Number of FGD and KII conducted (PDF)

S3 Table 3: Program performance assessment by evaluating the target vs achievement (PDF) S4 Figure 1: Demographic information of the respondents (PDF)

S5 Table 4: Results from the quantitative assessment (PDF)

S6 Figure 2: Nutritional status of the children whose caregivers or mothers attended SBC activities (Wasting)

## 11. Author Contributions

Conceptualization: Taslima Arzu, Md, Ariful Kabir Sujan, S.M. Symon Bappy & Md. Alamgir Hossain.

Data curation: Taslima Arzu, Md. Ariful Kabir Sujan, Md. Alamgir Hossain, Monowarul Islam, Viulon Prosad, Suparna Das Toma

Formal analysis: Taslima Arzu, Md. Ariful Kabir Sujan, Md. Alamgir Hossain, Monowarul Islam, Viulon Prosad Methodology: Taslima Arzu, Md. Ariful Kabir Sujan, Md. Alamgir Hossain

Software: Md. Ariful Kabir Sujan, Monowarul Islam, S, M, Symon Bappy Supervision: Taslima Arzu, Md. Ariful Kabir Sujan, Md. Alamgir Hossain Writing – original draft: Taslima Arzu, Md. Ariful Kabir Sujan

Writing – review & editing: Taslima Arzu, Md. Ariful Kabir Sujan, S.M. Symon Bappy, Md. Alamgir Hossain, Monowarul Islam, Suparna Das Toma, Vulon Prosad

