## Supplementary material for "Impact assessment of social behavioral change activities on infant and young child feeding (IYCF) of the nutrition program in the host community": Ethical Permission from the ethical authority

Government of the People's Republic of Bangladesh Office of the Civil Surgeon (CS) Office

Cox's Bazar

https://cs.coxsbazar.gov.bd/

No: 51.04.2200.009.16.22.24/03 Date: 24 September 2024

**Sub: Permission to conduct SBC study at the host community of Moheshkhali and Pekua**

Ref: WVB letter no. WVBCXBOM-15-09-24/01; Date- 15 September 2024.

In response to the letter mentioned above, this office is pleased to grant permission to the World Vision Bangladesh (WVB) to conduct "Impact assessment of social behavioral change activities on infant and young child feeding (IYCF) of the nutrition program in the host community, at Moheshkhali and Pekua of Cox's Bazar, under the following terms and conditions:

1. You are requested to inform and coordinate with the concerned Upazila Health and Family Planning Officer (UH&FPO) offices of Moheshkhali and Pekua before commencing the activities.
2. Government policy, rules, and laws must be strictly followed during the operation.
3. Survey questionnaires, contents, and final reports must be submitted to the CS office and respective UH&FPO offices before publication or dissemination.
4. This permission shall remain valid up to 31 December 2024.

24/09/24

Dr. Asif Ahmed Howlader
Civil Surgeon, Cox's Bazar


Taslima Arzu
Program Manager,
World Vision Bangladesh, Cox's Bazar

Copy for Information and necessary action:

1. Divisional Director (Health), Chattogram.
2. Upazila Nirbahi Officer (UNO), Moheshkhali and Pekua
3. Upazila Health and Family Planning Officer (UH&FPO), Moheshkhali and Pekua
4. Upazila Family Planning Officer (UFPO), Moheshkhali and Pekua
5. PS to CS (For the kind information of CS.
6. Office Copy.
