## Supplementary material for "Impact assessment of social behavioral change activities on infant and young child feeding (IYCF) of the nutrition program in the host community": Population type and quantitative sample for the assessment

**Table 1: Population type and quantitative sample for the assessment**

| Population type | Target | Sample |
|---|---|---|
| Pregnant and Breastfeeding women (PBW)<br>(Screening, counseling, MtMSG, messaging, etc.) | 15,221 | 224 |
| Child (0-59 months), but the respondent will be the mother and caregiver. (Screening, GMP, WBW, NAW) | 71,030 | 227 |
| Grand Total | 451 | |
