## Supplementary material for "Impact assessment of social behavioral change activities on infant and young child feeding (IYCF) of the nutrition program in the host community": Number of FGD and KII conducted

**Table 2: Number of FGD and KII conducted**

| What | Host | | Total |
|---|---|---|---|
| | Participant type/Activity | Quantity | |
| KII | Graduated MCG/MtMSG (graduated) | 3 | 8 |
| | Nutrition staff | 2 | |
| | GO/NGO relevant representative | 3 | |
| FGD | Cooking session of MtMSG, and local leader/Imam meeting. | 4 | 8 |
| | Caregivers of the under-five children | 4 | |
