## Supplementary material for "Impact assessment of social behavioral change activities on infant and young child feeding (IYCF) of the nutrition program in the host community": Program performance assessment by evaluating the target vs achievement

**Table 3: Program performance assessment by evaluating the target vs achievement**

| Indicators | Target | Achievement | % |
|---|---|---|---|
| Community MUAC Screening of U5 | 73,934 | 71,030 | 96% |
| Community MUAC Screening of PLW | 19,758 | 15,221 | 77% |
| Growth Monitoring and Promotion (GMP) | 81,163 | 71140 | 88% |
| IYCF group Messaging | 44,378 | 39,841 | 90% |
| Courtyard session at the cluster level | 5437 | 6192 | 114% |
| Mother to Mother Support Group (MtMSG) | 59 | 59 | 100% |
| Vitamin-A Campaign | 73934 | 93629 | 127% |
| Deworming Campaign | 49314 | 78,346 | 159% |
| IFA Supplementation for PBW with MIYCN messaging | 4057 | 3493 | 86% |
