## Supplementary figures and images for "Impact assessment of social behavioral change activities on infant and young child feeding (IYCF) of the nutrition program in the host community"

### Demographic information of the respondents (N=451)

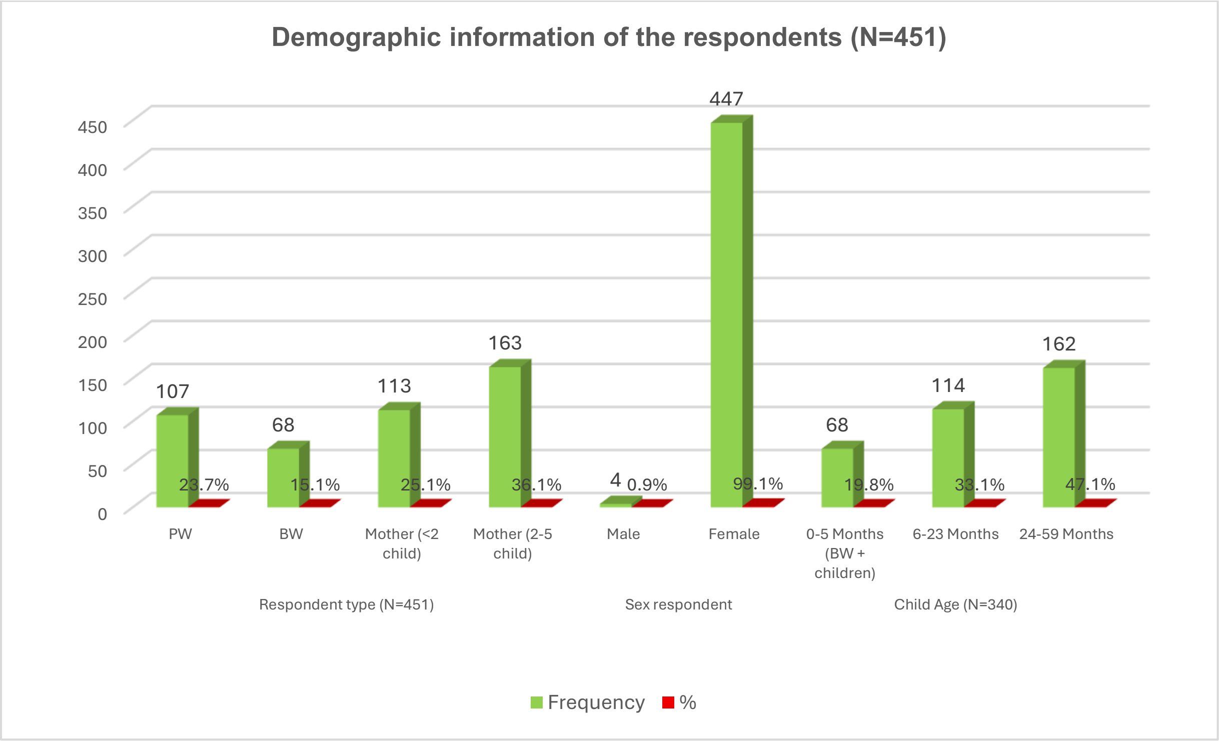

### Nutritional status of the children

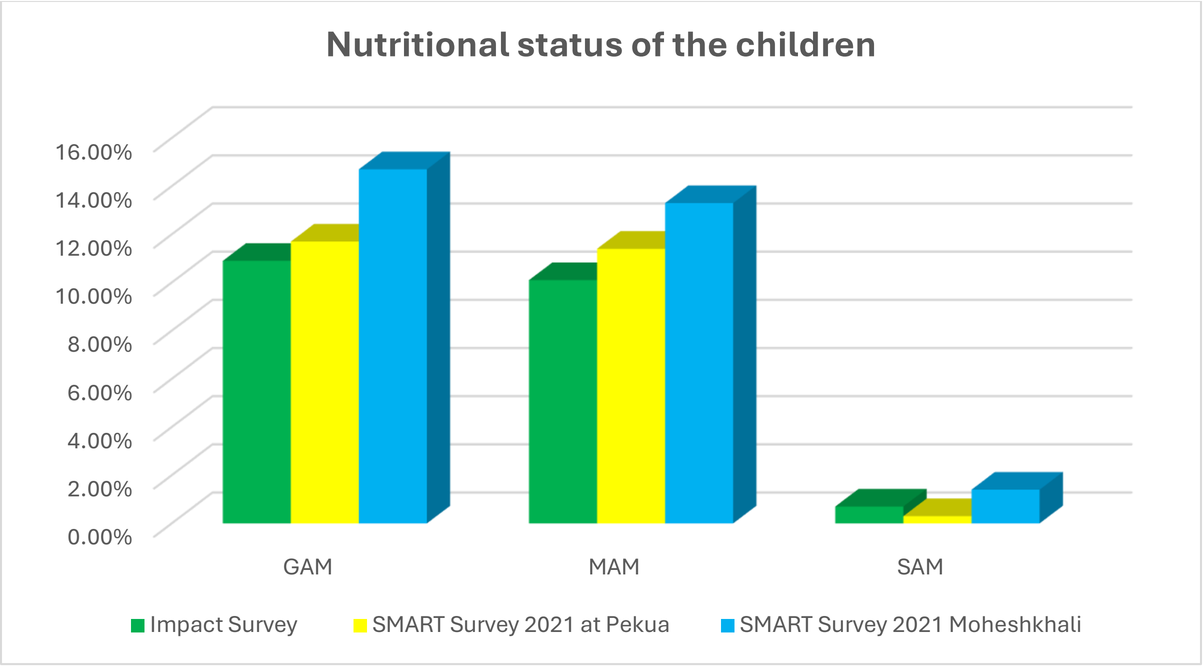
