## Supplementary material for "Impact assessment of social behavioral change activities on infant and young child feeding (IYCF) of the nutrition program in the host community": Results from the quantitative assessment

**Table 4: Results from the quantitative assessment**

| Indicators | Category | N | Response | % |
|---|---|---|---|---|
| Does CNV measure your child at the HH level | Yes | 451 | 416 | 92.2% |
| How many times did CNV visit HH for screening in the last quarter? | 1 time | 416 | 13 | 3.1% |
| | 2 time | | 17 | 4.1% |
| | 3 time | | 386 | 92.8% |
| Measured for GMP & Provide Counselling | Yes | 276 | 265 | 96.0% |
| Do you receive GMP key messages? | Yes | 276 | 261 | 94.6% |
| Did you receive the IYCF key message through MtMSG? | Yes | 289 | 262 | 90.7% |
| How many IYCF key messages are there? | 1-Message | 262 | 4 | 1.5% |
| | 2- Messages | | 11 | 4.2% |
| | 3-Messages | | 68 | 26.0% |
| | 4-Messages | | 179 | 68.3% |
| Did you receive one-to-one counseling? | Yes | 289 | 182 | 63.0% |
| Early Initiation of breastfeeding within 1 hour after delivery (aged 0-23 months) | | 308 | 301 | 97.7% |
| Exclusively breastfeeding (EBF) among children 0-5 months of age | | 115 | 84 | 73.0% |
| Children (aged 6–23 months) have Minimum Dietary Diversity (MDD) (at least 4 food groups) | | 193 | 102 | 52.8% |
| Minimum Acceptable Diet (MAD)for children 6-23 months | | 193 | 69 | 35.8% |
| In the past six months, have you changed any of your nutritional habits in a positive direction? | | 451 | 276 | 61.2% |
